# An Open-label Clinical Study of Brief Submaximal Cardiopulmonary Testing in Pre-surgical Evaluation: Feasibility of implementation

**DOI:** 10.1101/2024.01.24.24301611

**Authors:** Zyad J. Carr, Daniel Agarkov, Judy Li, Jean Charchaflieh, Andres Brenes-Bastos, Jonah Freund, Jill Zafar, Robert B. Schonberger, Paul Heerdt

**Affiliations:** Yale University School of Medicine, New Haven, Connecticut USA; Department of Anesthesiology, Yale New Haven Hospital, New Haven, Connecticut USA; Department of Anesthesiology, Bridgeport Hospital, Bridgeport, Connecticut USA

**Keywords:** Preoperative evaluation, submaximal cardiopulmonary exercise test, cardiac, pulmonary, evaluation, risk stratification

## Abstract

**Objectives:** We tested the logistic feasibility of integrating brief submaximal cardiopulmonary exercise testing (smCPET) in a pre-surgical evaluation (PSE) clinic.

**Design:** Prospective open-label clinical device trial.

**Setting:** Pre-surgical evaluation clinic.

**Participants:** 43 participants who met criteria of i) age ≥ 60 years old, ii) revised cardiac risk index of ≤2, iii) self-reported metabolic equivalents (METs) of ≥4.6 (i.e. ability to climb 2 flights of stairs), and iv) presenting for noncardiac surgery.

**Interventions:** Pre-intervention self-reported METs, Duke Activity Status Index (DASI) surveys, smCPET trial, Borg survey of perceived exertion, and post-intervention survey. **Measurements**: Feasibility endpoints were 1) operational efficiency as measured by length of time of experimental session ≤ 20 minutes, 2) no more than moderate perceived physical exertion as quantified by a modified Borg survey of perceived exertion of ≤7 in the absence of observed complications, 3) high participant satisfaction with smCPET task execution, represented as a score of ≥8, and 4) high patient satisfaction with scheduling of smCPET testing, represented as a score of ≥8.

**Results:** Session time was 16.9 minutes (±6.8). Post-test modified Borg survey was 5.35 (±1.8), corresponding to moderate perceived exertion. Satisfaction [on a scale of 1 (worst) to 10 (best)] regarding ease of smCPET tasks was 9.6 (±0.7) and mean patient satisfaction with smCPET scheduling was 9.5 (±1.5). Operational efficiency was achieved after 10-15 experimental sessions.

**Conclusions:** Our findings suggest that smCPET integration in a PSE clinic; 1) is time efficient 2) shows high participant satisfaction with task, and 3) rapidly achieved operational efficiency.

**Trial Registration:** ClinicalTrials.gov Registration: #NCT05743673. Principal Investigator: Zyad J. Carr, M.D. Date of Registration: 5-12-2023.

**Strengths and limitations of this study:**

- We examined patient-and logistic-centered acceptance of study procedures within the environment of a high-volume preoperative surgical evaluation clinic.
- Study procedures were well tolerated, and participants readily accepted submaximal cardiopulmonary exercise testing with high satisfaction with device use, scheduling, and perceived exertion.
- User operational efficiency developed over 10-15 sessions of use.
- This feasibility study met our proposed endpoints but is comprised of a small sample of participants, limiting its generalizability to larger populations.

## Background

Functional capacity or exercise tolerance, as measured by self-reported metabolic equivalents (METs), remains a cornerstone of preliminary assessment of fitness for surgery. METs are defined as multiples of the basal metabolic rate, conventionally defined as 3.5 ml.kg^-1^.min^-1^. Self-reported ability to climb one flight of stairs has a general consensus of 4 METs^1^. A threshold of <4.6 METs (self-reported inability to climb two flights of stairs) has been observed to correlate with major adverse cardiac events, all-cause mortality, and higher rates of perioperative complications^2–4^. However, self-reported and physician estimated METs remain insensitive in the accurate estimation of peak METs^5^ ^6^. Similarly, preoperative risk prediction tools are fragmented or have demonstrated significant limitations in capturing at-risk populations prior to surgical evaluation^7^. Thus, simple, reliable, and sensitive methods to improve the precision of preoperative evaluation continues to be an area of importance in preoperative assessment and the individualized identification of high-risk patients.

Traditional cardiopulmonary exercise testing (CPET) provides objective assessments of cardiopulmonary performance by analyzing measures of cellular respiration at rest and during exercise. Typically performed by measuring resting gas exchange followed by commencement of maximal exercise to expose pathophysiological impairments. CPET usually exploits a symptom-limited approach to stationary-cycle ergometer-derived exercise with a 3-minute resting stage, 3 minutes of unloaded cycling, and a 10-12 minute ramp stage with increasing resistance until terminated by the participant^8^. Abnormalities have been shown to be associated with perioperative morbidity after noncardiac surgery^9^. A peak VO_2_ of <15ml/kg/min has been frequently reported in the literature as a threshold for elevated perioperative cardiopulmonary complications in patients after thoracic and major noncardiac surgery^10–13^. In addition, CPET-derived peak VO_2_ has been observed to predict surgical site infection, postoperative respiratory failure, and increased risk of critical care readmission but not 30-day mortality and non-fatal myocardial infarction^14^. Despite its prognostic value for perioperative complications, traditional CPET has been limited in its widespread adoption for preoperative evaluation due to limited availability, required technical skills, necessity of maximal patient effort, complexity of task, and cost.

In contrast to traditional CPET, submaximal cardiopulmonary exercise test (smCPET) utilizes graded exercise and concomitant gas exchange analysis to provide a granular and personalized assessment of cardiopulmonary performance^8^. Several advantages are provided by smCPET over traditional CPET. First, a submaximal exercise effort is required since it analyzes the oxygen uptake efficiency slope (OUES) to extrapolate reliable estimates of peak METs and peak VO_2_^15–17^. The OUES predictive capability allows effort-independent estimation of extrapolated peak cardiopulmonary functional reserve, a particular advantage in deconditioned, frail, or functionally limited patient populations. Furthermore, as a time-limited assessment, smCPET may be efficiently integrated into conventional clinic schedules. Lastly, new devices have continued to miniaturize the smCPET footprint, permitting easy storage and transport. smCPET has demonstrated reliable prediction of length of stay and prediction of postoperative complications after noncardiac surgery^18^.

Despite these advantages, widespread adoption of smCPET for the purposes of preoperative evaluation has not been observed. Thus, it is unclear if smCPET can be feasibly integrated into a high-volume pre-surgical evaluation clinic setting.

### Study Objectives

We examined the feasibility of integration of brief smCPET into a high-volume pre-surgical evaluation (PSE) clinic of a large quaternary care facility. This initial study stage was performed for the purpose of determining the adequacy of study and patient-centered processes of the primary observational study which will examine the relationship of smCPET and perioperative outcomes, as measured by the postoperative morbidity survey (POMS)^19^. Our measured feasibility endpoints were 1) operational efficiency as measured by length of time of experimental session ≤ 20 minutes, 2) no more than moderate perceived physical exertion as quantified by a modified Borg survey of perceived exertion of ≤7 in the absence of observed complications, 3) high participant satisfaction with smCPET task execution, represented as a score of ≥8, and 4) high patient satisfaction with scheduling of smCPET testing, represented as a score of ≥8.

A prior study examining CPET and subjective clinician estimation had a sensitivity of 19.2% in the identification of patients with low functional capacity (≤4 METs)^14^. We were interested in quantifying if this was also present in our feasibility cohort using smCPET equivalents. Secondary outcomes included a comparison of differences between 1) self-reported METs survey vs. smCPET equivalent (extrapolated peak METs), 2) Duke Activity Status Index^20^ (DASI) vs. smCPET equivalent (extrapolated peak METs) and 3) estimated DASI maximal oxygen consumption (estimated peak VO_2_) vs. smCPET equivalent (extrapolated peak VO_2_).

## Materials and Methods

### Study Design

This is an ongoing prospective open-label clinical device trial approved by the Yale University Institutional Review Board (IRB#2000033885; ClinicalTrials.gov Registry. #NCT05743673. Principal Investigator: Zyad J. Carr, M.D. Date of Registration: 5-12-2023). **Study Population:** We successfully enrolled 43 participants who met the inclusion criteria of age ≥ 60 years old, with a revised cardiac risk index^21^ of ≤2, and self-endorsed subjective metabolic equivalents of ≥4, presenting for moderate to high-risk noncardiac surgery. The aim was to recruit 40 participants for this initial feasibility stage of the study. We estimated this number to be adequate to identify any study-related logistic process problems, patient-centered outcome deficiencies, and to determine operational efficiency.

We pre-screened by chart review and excluded potential participants with recorded severe or critical heart valve disease, active exertional angina, non-ambulation, gait abnormalities, end-stage renal disease, severe peripheral vascular disease, and neurological motor deficits. We excluded non-English speaking participants, those under legal guardianship, and participants documented to not have personal health care decision-making capacity. After pre-screening, a phone call was placed by a study team member to the potential participant, and eligible participants were invited for in-person informed consent, preoperative evaluation, questionnaire assessment of METs, and a smCPET experimental session.

### Testing Environment

Testing was performed at the PSE Clinic at Yale New Haven Hospital which is typically responsible for approximately >40,000 preoperative evaluations per year. On a daily basis, the PSE clinic is staffed by an anesthesiologist, 2 resident physicians, 3 certified nurse practitioners and 6 nursing staff and contains six exam rooms.

### Study Apparatus

The FDA-approved Shape II® system is a compact cardiopulmonary breath by breath exercise testing system that uses sub-maximal exercise effort to generate multiple quantitative measures of actual and extrapolated peak exercise tolerance. The device has been previously validated to conventional CPET measurements^22^. The compact design allows all the necessary equipment to be placed on a standard rolling cart and it was deployed in a PSE clinic examination room (2.4 x 2.4 meters). The device requires 2 minutes of baseline data, 3 minutes of escalating exercise using a stationary step and 1 minute of recovery data to generate a variety of individual measures of cardiac and pulmonary physiological data (**Supplementary Table 1**).

### Study procedures

On presentation to the PSE clinic, participants received height/weight and vital sign measurements (heart rate, blood pressure and pulse oximetry). Informed consent was performed, and participants were instructed on smCPET testing (∼5 minutes). Session time was measured from the beginning of pre-test METs questionnaires until the termination of the smCPET recovery phase. Study pre-test instruments included a self-reported 7-question METs assessment and the 12-question DASI survey. A post-test modified Borg survey of perceived exertion was performed after smCPET session and was recorded immediately after termination of the smCPET trial. After study interventions, a standard preoperative evaluation was completed, and the participant was discharged. A 24-hour post-experiment survey of minor/major complication and patient satisfaction was performed by telephone (**Supplementary Table 2**). With the exception of the patient satisfaction survey, all survey instruments were adapted from prior publications^23–25^. DASI peak METs and peak VO_2_ was calculated from individual participants DASI score using the recommended formula.

### Data analysis

Continuous variables are described as mean (standard deviation), ordinal variables as median (range), and categorical variables as number (percent).

## Results

### Participant recruitment

Participants were recruited from June 2023 through October 2023. We identified 209 potential participants that met eligibility criteria, 6 did not meet inclusion criteria, 59 failed pre-screening criteria and 89 declined study participation (**Figure 2**). Initially 46 participants were enrolled but 3 were excluded (operator error: 2; surgery cancellation: 1) for a final cohort of 43 participants.

**Figure 1.** A visual representation of the smCPET device in the PSE Clinic. (Model: co-author JF)

**Figure 2.**
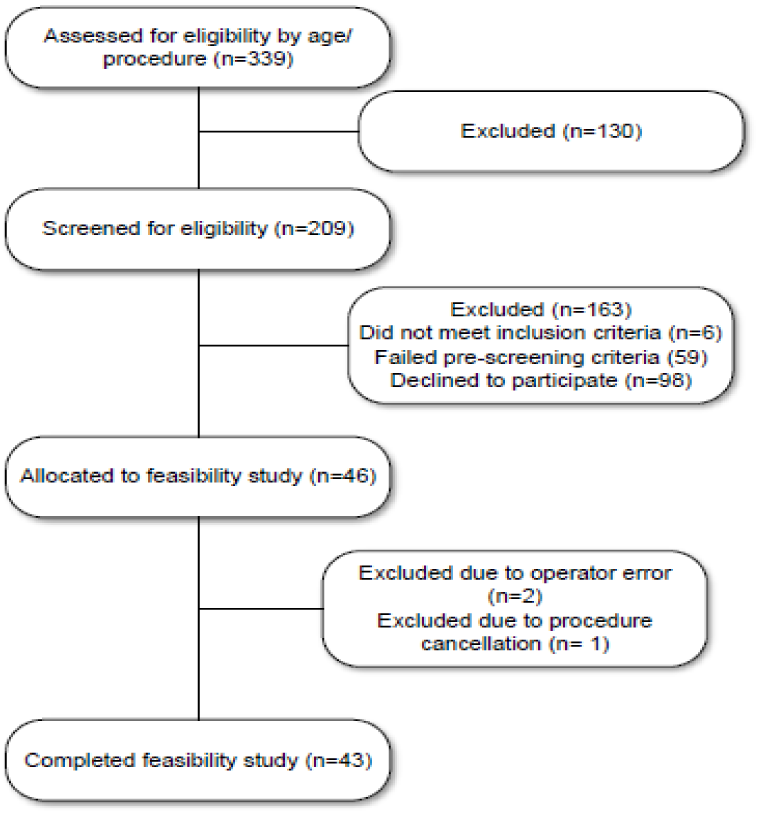
A Flow Diagram of Participant Enrollment.

### Baseline characteristics

Trial participants had a median age of 68 (range: 60-86 years old), 46.5% were female, and mean body mass index (BMI) was 27.5 (±6.0 kg/m^2^). Preoperative RCRI was a median of 1 (range: 1-2). Essential hypertension (51.2%), hyperlipidemia (39.5%) and solid tumor (58.1%) were the most common pre-morbid conditions. Former or active smokers comprised 51.2% of the cohort. Major abdominal surgeries (62.8%) comprised the majority of the noncardiac surgical procedures. **Table 1** describes the baseline demographics of the study population.

**Table 1.** Baseline Demographical Data of the Study Cohort (n=43).

| Table 1. Baseline Demographical Data of the Study Cohort (n=43) |  |  |
| --- | --- | --- |
| Age, in years, median (range) | 68 | (60-86) |
| Gender |  |  |
| Male | 23 | (53.5%) |
| Female | 20 | (46.5%) |
| Body Mass Index, in m/kg<sup>2</sup>, mean (SD) | 27.5 | (±6.0) |

| Revised Cardiac Risk Index Score, median (Range) | 1 | (1-2) |
| --- | --- | --- |
| **Preoperative Comorbidities** |  |  |
| Essential Hypertension | 22 | (51.2%) |
| Hyperlipidemia | 17 | (39.5%) |
| Ventricular Dysrhythmia | 1 | (2.3%) |
| Congestive Heart Failure | 1 | (2.3%) |
| Myocardial Infarction | 3 | (7.0%) |
| Cerebrovascular Disease | 1 | (2.3%) |
| Chronic Obstructive Pulmonary Disease | 3 | (7.0%) |
| Asthma | 4 | (9.3%) |
| Obstructive Sleep Apnea | 3 | (7.0%) |
| History of Prior Lung Resection | 1 | (2.3%) |
| Diabetes Mellitus | 7 | (16.3%) |
| Thyroid Disorders | 7 | (16.3%) |
| Solid Tumor | 25 | (58.1%) |
| Anemia | 1 | (2.3%) |
| **Social History** |  |  |
| **Smoking** |  |  |
| Active | 4 | (9.3%) |
| Former | 18 | (41.9%) |
| Never | 21 | (48.8%) |
| Marijuana Use (active) | 4 | (9.3%) |
| **Alcohol Use** |  |  |
| Active | 24 | (55.8%) |
| Former | 16 | (37.2%) |
| Never | 3 | (7.0%) |
| **Cardiovascular Medication Use** |  |  |
| Beta-blocker | 14 | (32.6%) |
| Calcium channel antagonist | 9 | (20.9%) |
| ACE/ARB antagonist | 16 | (37.2%) |
| Diuretic | 12 | (27.9%) |
| **Surgical Categories** |  |  |
| Abdominal Major | 27 | (62.8%) |
| Musculoskeletal Major | 4 | (9.3%) |
| Neurosurgical Major | 2 | (4.7%) |
| Thoracic Major | 5 | (11.6%) |
| Other Major | 5 | (11.6%) |

### Feasibility and participant smCPET acceptability

The mean (SD) experimental session time was 16.9 minutes (±6.8). The mean (SD) modified Borg survey after experimental sessions was 5.35 (±1.8), corresponding to moderate perceived exertion. On 24-hour post-experimental session survey, a total of 43 (100%) of participants were reached. Mean (SD) patient satisfaction [on a scale of 1 (worst) to 10 (best)] was 9.5 (±1.5). The mean (SD) ease of performing smCPET tasks was reported as 9.6 (±0.7). Among this cohort, no major or minor complications associated with study testing were reported by participants (0/43; 0%). Operational efficiency was achieved within 10-15 experimental session among four study team members who trained on the device.

### Secondary measures

Average self-reported peak METs was higher when compared to smCPET equivalent (extrapolated peak METs) [7.6 (±2.0) vs. 6.7 (±1.8)]. DASI estimated peak METs was higher when compared to smCPET equivalent (extrapolated peak METs) [8.8 (±1.2) vs. 6.7 (±1.8)]. DASI-estimated peak VO_2_ was higher than smCPET equivalent (extrapolated peak VO2) [30.9 (±4.3) vs. 23.6 (±6.5)]. **Figure 3** provides a comparison of values obtained from smCPET compared to self-reported peak METs, DASI peak METs, and peak VO2.

**Figure 3.**
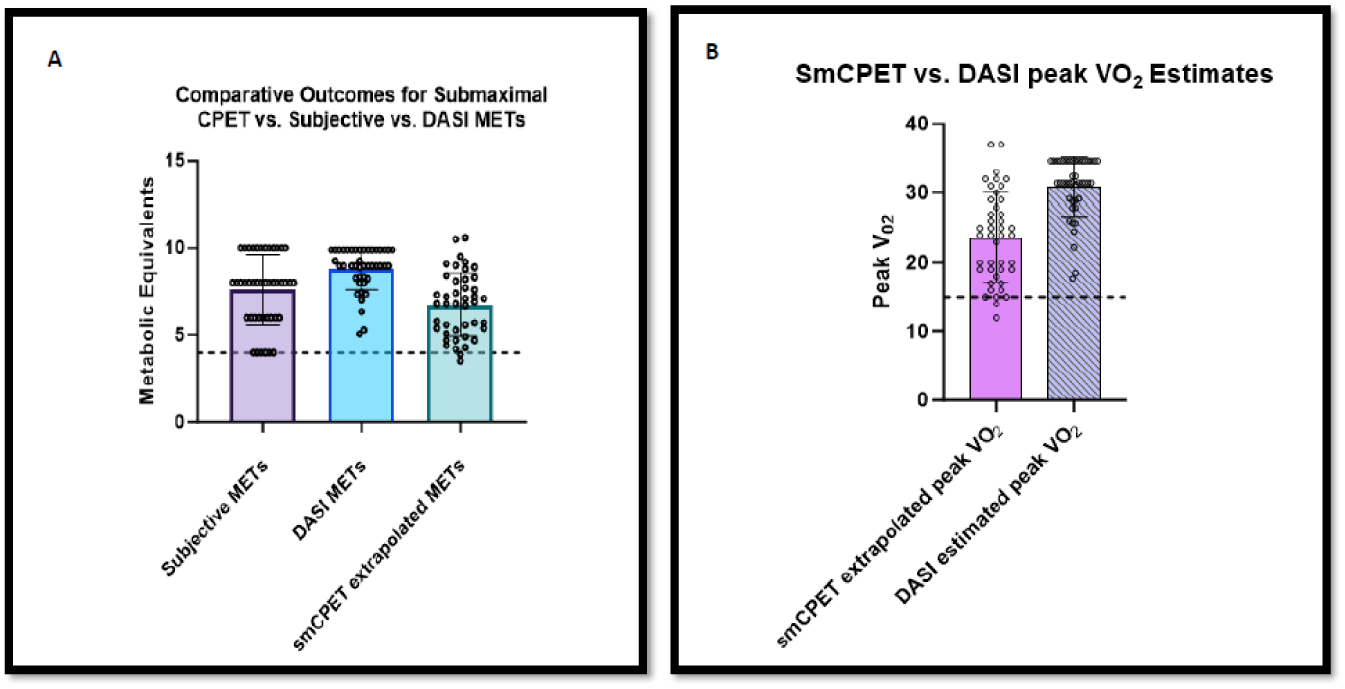
Figure 3a reports differences between estimated peak METs between self-reported and extrapolated peak METs derived from smCPET. Figure 3b reports differences between Duke Activity Status Index estimated peak VO_2_ and submaximal CPET extrapolated peak VO_2_.

## Discussion

The integration of brief smCPET in a high-volume PSE clinic is feasible as measured by endpoints of session time, patient satisfaction with smCPET task execution, perceived exertion, and session scheduling. Secondly, operational efficiency of study team members was acceptable within 10-15 experimental sessions. Lastly, we observed consistent underestimation of self-reported METs, DASI peak METs, and DASI peak VO_2_ when compared to smCPET equivalent values. We found that smCPET set-up, calibration, patient instruction, and execution of the study trial was time efficient. Mean session time was 16.9 minutes with rapid improvement over the study time period as operators (n=4) became facile with the study instrument (**Figure 4**). In fast paced, high-volume clinic environments, this efficiency is important, as patients are often seen short notice for preoperative evaluation. Given the short time requirement, we were able to flexibly arrange smCPET testing around other clinic appointments, facilitating successful study recruitment, and decreasing time burden on participants. PSE clinic-performed smCPET was also associated with a high level of patient satisfaction related to ease of task performance, and perceived exertion. The tested device uses a stair-step for graded exercise, which was often familiar to participants. The short duration of graded exercise, with automated verbal prompts to increase work rate by the device, was not perceived by any participant as maximum effort by Borg survey. No exercise-related major or minor complications were observed, and patients were consistently encouraged to safely provide maximal effort within the graded exercise portion of smCPET. Early termination of conventional CPET trials, due to participant fatigue or safety considerations, has been reported to be approximately 11%, no participant in our feasibility cohort elected early trial termination^14^. It is important to note that we selected for functionally independent participants with self-reported >4.6 METs, as represented by the ability to climb two flights of stairs, and expansion to less functional patients may result in higher failure rates. However, smCPET has been successfully tested in high-risk and frail populations, suggesting that a wide spectrum of preoperative populations could be tested using smCPET^26–28^.

**Figure 4.**
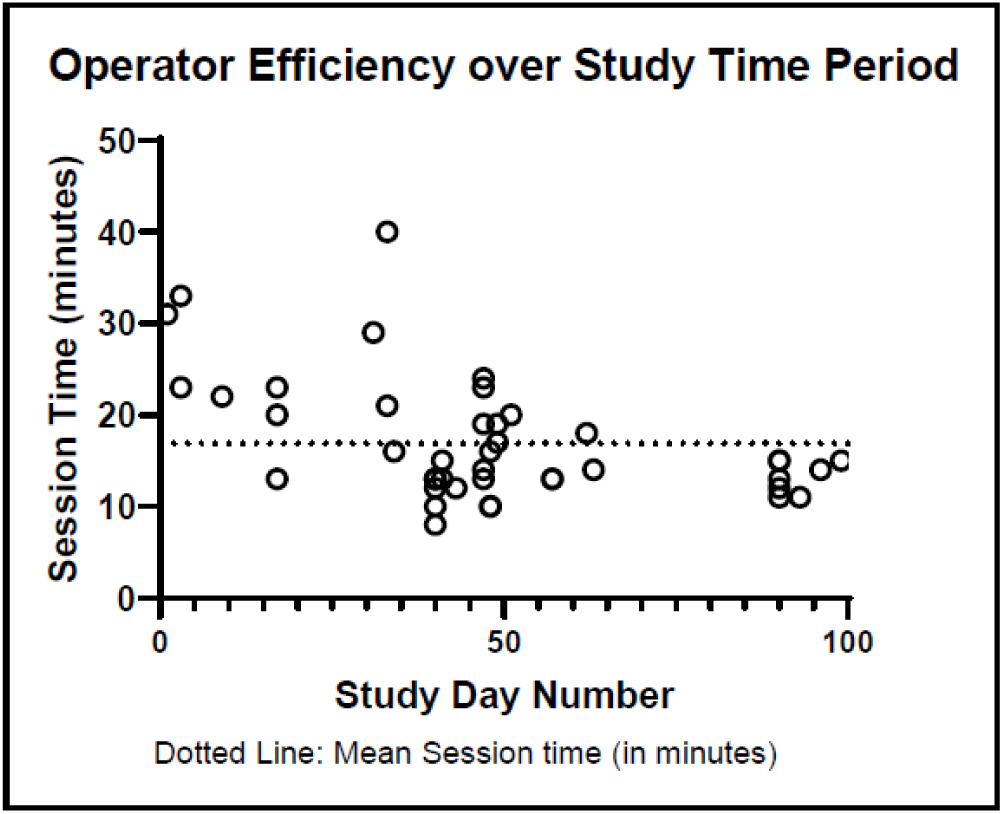
Operator Efficiency Measured by Session Time over the Study Time Period.

In prior work, Wijeysundera and colleagues^14^ observed that subjective clinician estimation had a sensitivity of 19.2% in the identification of patients with low functional capacity (<4 METs)^14^. We observed that self-reported subjective METs and DASI estimated METs were, on average, lower than their smCPET equivalent (extrapolated peak METs). In our analysis, 18.6% (N=8/43) of participants self-reported their peak METs within 10% of smCPET extrapolated peak METs, 27.9% of participants underestimated (N=12/43) and 53% (23/43) overestimated their peak METs by >10%, respectively. smCPET identified that 18.6% (n=8) of our study cohort had ≤4.7 extrapolated peak METs, correlating very closely to a METs threshold associated with higher perioperative cardiovascular morbidity and mortality. Furthermore, smCPET identified that 20.9% of our cohort had an age adjusted peak VO_2_ of <20ml/kg/min, corresponding to poor aerobic capacity, and 4.6% of our cohort achieved an extrapolated peak VO_2_ <15 ml/kg/min. It has been shown that conventional preoperative evaluation may not improve perioperative outcomes^29^. This may suggest a useful role for more precise risk stratification using brief smCPET in preoperative testing. Despite supporting evidence, widespread adoption of CPET and new generation smCPET devices has not been observed in preoperative testing. This is likely multifactorial due to limited awareness of new generation smCPET devices, perceived overhead cost, perceived time constraints, perceived operational complexity, and lack of clinical evidence regarding preoperative clinic integration. However, further knowledge of smCPET predictive validity and optimal system processes for selecting patients is required to identify its preoperative testing indications and its role in preoperative evaluation.

### Study Limitations

This study had several limitations related to generalizability to other populations. As an open-label device clinical trial using a convenience sample of preoperative patients, we deliberately excluded patients with high levels of comorbid conditions as quantified by RCRI. Although we are not able to generalize to this population, our goal was to establish feasibility of brief smCPET assessment within a presumed healthy but older perioperative cohort that would have likely not been captured by extensive preoperative evaluation. Secondly, although published data has validated smCPET predictive performance with perioperative cardiovascular morbidity and mortality, our cohort is not yet powered for assessment of these outcomes. Finally, our demonstration of no device-related adverse events is reassuring, but it should be cautiously interpreted given the small sample size and possibility of rare exercise-induced adverse events.

## Conclusions

In summary, we observed that smCPET was well accepted into the workflow of a high-volume PSE clinic. All logistical, operational, and patient-centered feasibility endpoints were met. Operator efficiency with the smCPET instrument was rapid and achieved relative parity at Day 30 or 10-15 sessions. This feasibility analysis has, 1) reinforced the structural integrity of our active study protocol assessing relationships of smCPET findings with perioperative outcomes, 2) affirmed satisfactory patient-centered outcomes with the study procedures, and 3) provided insight into functional capacity variation in a cohort of older, but otherwise functionally independent, adult participants. Further studies should examine smCPET predictive validity and optimal system processes for patient selection.

## Statements & Declarations

### Author Contributions

Study conception and design were performed by Zyad J. Carr, Paul Heerdt, Robert B. Schonberger. The first draft of the manuscript was performed by Zyad J. Carr, Jean Charchaflieh, Jill Zafar, Andres Brenes-Bastos. Data collection was performed by Zyad J. Carr, Jonah Freund, Judy Li, Daniel Agarkov. Statistical analysis was performed by Zyad J. Carr. Further manuscript editing was performed by all co-authors. All authors read and approved the final manuscript.

### Ethics Approval

This study was performed in accordance with the principles of the Declaration of Helsinki. Approval was granted by the Yale University (New Haven, Connecticut, USA) Institutional Review Board (IRB#2000033885; ClinicalTrials.gov Registry. #NCT05743673.

Principal Investigator: Zyad J. Carr, M.D. Date of Registration: 5-12-2023).

### Consent to participate

Informed consent was obtained from all individual participants included in the study.

### Consent to publish

All participant data is deidentified. Co-author Jonah Freund has given express written consent for publication of his image in Figure 1.

## Data Availability

All data produced in the present study are available upon reasonable request to the authors

## Acknowledgements

We would like to acknowledge the work of Rayna Lewoc, DNP, APRN, CRNA, Elizabeth Womack, Elizabeth Cozza, APRN, Gwendolyn Burkett, and the staff of the Yale-New Haven Hospital Pre-Surgical Evaluation Clinic.

## Patient and Public Involvement

Patient or the public were not involved in the design, or conduct, or reporting, or dissemination plans of our research.

---

**Supplementary Table 1.** Summary and Selected Measurements of Submaximal Cardiopulmonary Exercise Testing.

| Variable | Description | Commentary |
| --- | --- | --- |
| **Summary of Shape II® metabolic analyzer used in study:** The device uses breath by breath sampling during calibration and exercise challenge using a differential pressure pneumotach method for volume calibration and measurement, an infrared sensor for CO<sub>2</sub> and a paramagnetic sensor for O<sub>2</sub> measurements. Automated calibration using a calibration gas mixture (15.6% O<sub>2</sub>/5% CO<sub>2</sub>) is performed at regular intervals. The Shape II calculations used for causes of exertional dyspnea differentiation are Artificial Intelligence (AI) based algorithms previously calibrated and validated against conventional cardiopulmonary exercise testing methods. |  |  |
| HR (resting) | Resting heart rate | Measured during 1st stage prior to exercise. |
| HR (peak) | Peak heart rate achieved during exercise | Measured during 2nd stage during exercise. |
| % HR reserve utilized | Percentage of heart rate reserve utilized | Percentage of resting heart rate and the age-dependent predicted maximum heart rate |
| % HR max predicted attained | Percentage of predicted maximum achieved heart rate achieved | Difference between maximum measured heart rate during exercise and age-dependent estimated maximum heart rate. |
| CRI | Chronotropic Recovery Index | A measure of heart rate recovery after exercise |
| RR rest | Respiratory rate at rest | Measured during 1st stage prior to exercise. |
| RR end exercise | End-exercise respiratory rate | Measured during 2nd stage during exercise. |
| End-tidal CO<sub>2</sub> (rest) | End-tidal carbon dioxide, at rest | Influenced by cardiac output, pulmonary vascular resistance, and chronic hypoventilation syndromes |
| End-tidal CO<sub>2</sub> (peak) | End-tidal carbon dioxide, peak exercise | Peak carbon dioxide during exercise |
| Resting SpO<sub>2</sub> | Resting pulse oximetry | Resting peripheral oxygenation |
| Peak SpO<sub>2</sub> | Peak exercise pulse oximetry | Peak exercise peripheral oxygenation |
| RER | Respiratory Exchange Ratio | Ratio between metabolic production of CO<sub>2</sub> and uptake of O<sub>2</sub> |
| VE/VCO<sub>2</sub> slope | Minute ventilation/CO<sub>2</sub> production slope | Breathing efficiency slope reflects the efficiency of elimination of CO<sub>2</sub> |
| Δ EtCO<sub>2</sub> (rest to end exercise) | Change in end-tidal carbon dioxide during exercise | A measure of cardiac output and pulmonary blood flow |
| Gxcap (peak) | Gas exchange-derived pulmonary vascular capacitance at peak exercise | Shown to correlate with cardiac output and inversely with pulmonary vascular resistance (PVR) and DLCO; in patients with PAH or PVH, Gxcap is reduced due to increased PVR. |
| OUES (linear slope) | Oxygen uptake efficiency slope, percentage of expected | Assesses how well oxygen is extracted from the inhaled air, distributed to the muscles by the cardiopulmonary system, and utilized by the muscles in energy metabolism; Particularly valuable in assessing exercise capacity and response to medications or rehabilitation |
| Peak Attained METs | Metabolic Equivalents | Metabolic equivalents attained during submaximal exercise. |
| Peak Extrapolated METs | Metabolic Equivalents | Peak estimated metabolic equivalents |
| O<sub>2</sub> pulse | Amount of oxygen consumed per heartbeat |  |
| Sub-maximal VO<sub>2</sub> (peak attained) | Sub-maximal exercise oxygen uptake | Achieved peak maximal oxygen uptake during submaximal exercise. |
| Extrapolated maximum VO<sub>2</sub> | Predicted maximal exercise oxygen uptake | Calculated peak maximal oxygen uptake. |
| MVI score | Dimensionless; (cumulative sum score) | Combined cardiopulmonary index with threshold value of normal vs. impairment. |
| Cardiac disease silo score | Dimensionless; (cumulative sum score) | Parameters: VE/VCO<sub>2</sub> slope, O<sub>2</sub> pulse to VO<sub>2</sub> slope, circulatory equivalents, HR recovery |
| Pulmonary Vascular Disease silo score | Dimensionless; (cumulative sum score) | Parameters: VE/VCO<sub>2</sub> slope, peak Gxcap, resting SpO<sub>2</sub>, SpO<sub>2</sub> desaturation |
| Chronic Obstructive Pulmonary Disease silo score | Dimensionless; (cumulative sum score) | Parameters: FEV1 % Predicted, breathing reserve, SpO<sub>2</sub> desaturation, P mixed expired CO<sub>2</sub>/P end-tidal CO<sub>2</sub> ratio (V/Q ratio) |
| Restrictive Lung Disease silo score | Dimensionless; (cumulative sum score) | Parameters: FVC % Predicted, SpO<sub>2</sub> desaturation, VT max / VT rest, RR/VCO<sub>2</sub> slope (lung stiffness) |
| De-conditioning silo score | Dimensionless; (cumulative sum score) | Parameters: extrapolated peak VO<sub>2</sub>, % ideal BMI, HR to VO<sub>2</sub> linear regression slope, HR recovery 1 minute post exercise |
| V/Q plot | Dimensionless; (cumulative sum score); (Normal, Left ventricular dysfunction, Chronic Obstructive Pulmonary Disease, Transitional or Pulmonary Arterial Hypertension) | Assesses which physiological area may contribute to visualized impairments during submaximal exercise testing. |
| AwCap peak (VT pk x PECO<sub>2</sub> peak) | airway capacitance; L x mmHg | Airway Capacitance |
| VT pk (peak attained VT) | Peak attained tidal volume in L/min | Value obtained during exercise. |
| VE peak (peak attained VE) | Peak attained minute ventilation in L/min | Value obtained during exercise. |

Abbreviations: HR; heart rate; CRI; chronotropic Recovery Index; RR; respiratory rate (in breaths/min); CO<sub>2</sub>; carbon dioxide; SpO<sub>2</sub>; pulse oximetry; RER; respiratory Exchange Ratio; Gxcap; pulmonary arterial capacitance; OUES; oxygen uptake efficiency slope; METs; metabolic equivalents; VO<sub>2</sub>; oxygen consumption; MVI; multivariable index; VE; minute ventilation; VT; tidal volume; AwCap; airway capacitance.

**Supplementary Table 2.** Adapted Subjective METs Survey.

| **Supplementary Table 2. Adapted Self-Reported Subjective Metabolic Equivalents Survey** |  |  |
| --- | --- | --- |
| Question | Estimated Metabolic Equivalents | Accepted value |
| *Can you perform the following activities (yes/no)* |  |  |
| Watching television, writing, desk work? | 1-2 | 2 |
| Walk slowly on level ground (1.7mph) | 2-3 | 3 |
| Climb two flights of stairs, without stopping to rest? | 3-5 | 4 |
| Walk at moderate pace on level ground (3mph/20 minute mile), ride a stationary bicycle at very light intensity or vacuum around the home? | 3-5 | 4 |
| Ride a stationary bicycle at moderate intensity? | 5-6 | 6 |
| Jogging, fast swimming, play soccer or tennis? | 7-8 | 8 |
| Run a 7.5 minute/mile, jump-rope 100 skips/minute, run up the stairs | 10-13 | 10 |

